# Two Stage Hierarchical Group Testing Strategy to Increase SARS-CoV-2 Testing Capacity at an Institution of Higher Education: A Retrospective Analysis

**DOI:** 10.1101/2021.04.14.21255494

**Authors:** Troy Ganz, Markus Waithe-Alleyne, Deirdre Slate, Rachel Donner, Kevin Hines, Gyorgy Abel, Jared Auclair

## Abstract

Population testing for severe acute respiratory syndrome 2 (SAR-CoV-2) is necessary owing to the possibility of viral transmission from asymptomatic cases, yet scarcity of reagents and equipment has added to the cost-prohibitive implementation of screening campaigns at institutions of higher education. The high analytical sensitivities of leading nucleic acid amplification diagnostic methods allow for group testing to increase testing capacity. A feasibility study was performed using an optimized testing configuration model for pooling three, five, and ten samples. Following the standard RNA extraction and purification workflow for quantitative reverse transcriptase polymerase chain reaction (RT-qPCR) method using Thermo Fisher TaqPath™ COVID-19 multiplex primers and probes for the ORF1ab, N, and S genes, matrix and dilution effects were assessed using pooled negative samples as the diluent. Probit analysis produced a limit of detection of 16075 (ORF1ab), 1308 (N), and 1180182 (S) genomic copy equivalents per milliliter. Trials comparing neat to 1:5 dilution for 34 weak-to-strongly positive samples demonstrated average threshold cycle (CT) shifts of 2.31±1.16 (ORF1ab), 2.23±1.12 (N), and 2.79±1.40 (S). Notwithstanding observed S gene dropouts, the false negative rate was unaffected. As the ratio of asymptomatic positive to symptomatic positive SARS-CoV-2 infected individuals was approximately 4:1 and the average prevalence was 0.16% since we started testing in August 2020, pooled testing was identified as a viable, cost-effective option for monitoring the Northeastern University community.

## Introduction

The emerging pandemic of coronavirus disease (COVID-19), sourced from the novel strain of beta-coronavirus known as Severe Acute Respiratory Syndrome Coronavirus 2 (SARS-CoV-2), has had a severe impact on global healthcare systems [1,2]. Institutions of higher education (IHE) have been confronted with the conflicting obligations to guarantee a standard quality of education while ensuring the safety of their faculty, staff, and students during the pandemic. For universities with shared living, dining, transportation, and classroom learning arrangements, the greater requirements of risk management must be weighed against the economic loss of closing for an indeterminate period [3,4]. Further pressures are imposed upon IHE in urban environments as positive correlations of transmission have been observed with factors such as reduced air quality and higher population density [5]. As with other respiratory infections, transmission is facilitated by viral shedding in the upper respiratory tract that may release viral particles in the form of aerosols (≤5 µm) or droplets (>5 µm) to contaminate the surrounding air or surfaces, and spread via fomites, coughing, sneezing, and exhalation from symptomatic, as well as pre-symptomatic and asymptomatic, carriers [6]. Along with recommended measures for risk management such as physical distancing, obligatory personal hygiene, and mask wearing, a rigorous screening and surveillance campaign provides the most effective response.

Quantitative reverse transcriptase polymerase chain reaction (RT-qPCR) is the most sensitive method for early detection of SARS-CoV-2 when compared to other available analytical methodologies (e.g., lateral flow immunoassay), which require larger viral titers or longer periods of seroconversion following infection [7,8]. Nasopharyngeal, nasal (nares), or oropharyngeal respiratory samples, and saliva have been evaluated through several RT-qPCR platforms; the added benefit being upper respiratory collections are less invasive than venipuncture for serological testing, resulting in better patient compliance [8]. Drawbacks coincide with the sharp increase in global demand for medical and testing supplies such as personal protective equipment (PPE; gloves, gowns, surgical and N95 masks), collection swabs and containers, transport and lysis buffers, nucleic acid extraction and amplification kits, consumables such as micropipette tips, analytical equipment, as well as qualified technical personnel as the high-complexity format of RT-qPCR testing schema requires a reliable supply chain of critical materials.

Group testing, or sample pooling, is an attractive method to increase testing capacity without a need for additional resources or training. Since the introduction of the methodology by Dorfman in 1943, subsequent studies have generated models to predict optimal group sizes within a test’s analytical sensitivity and relative cost as factors of dilution and individual repeat testing, or deconvolution, must be considered [10,11]. Pressured by the COVID-19 pandemic, investigators have sought to validate SARS-CoV-2 group testing on available molecular platforms [12,13,14,15]. The following investigation sought to define an ideal group size with respect to the cost of the current protocol while retaining an acceptable analytical sensitivity.

## Methods

### Sample Collection

The study was conducted at the Life Science Testing Center (LSTC; Burlington, MA, USA) for Northeastern University (NEU) students, faculty, and staff within the United States Northeast (MA, ME) regional campuses. Population surveillance includes sample collection from symptomatic, pre-symptomatic, asymptomatic, and uninfected individuals.

Anterior nasal swabs were collected in 3 mL BD Vacutainer® (without additives) tubes and transported dry at ambient temperature. Processing involved addition of 3 mL viral transport medium (VTM; Redoxica, Little Rock, AR, USA) and shaking at 1000 rpm for 5 minutes. Specimens are stable for <72 hours at 2-8°C and indefinitely at - 80°C.

### Modeling

Our prospective workflow can be modeled using an adaptive two-stage hierarchical algorithm, whereby results from the master pool direct repeat testing of the individual samples. An optimal testing configuration is a result of several parameters including disease prevalence, group size, and analytical sensitivity and specificity. Significant to our protocol, success of the MS2 phage extraction control defines an additional retesting criterion that may be interpreted to impact the master pool. Utilizing a web-based R application (www.chrisbilder.com/shiny/), a variable was included to reflect the probability of MS2 failure (invalid) to achieve estimated reductions for pools of size 3, 5, and 10. Sensitivity was set to 95% based on the calculated limit of detection, while specificity was set to 99% under the combined influence of the MS2 control and operator review. Results were depicted to distinguish the number of tests required for 1000 samples as informed by both the Shiny app and the modified version for each group size with respective invalid rates and prevalence using Excel.

### RNA Extraction and RT-qPCR

SARS-CoV-2 RNA was extracted from 200 µL of sample using the MagMAX™ Viral/Pathogen Nucleic Acid Isolation Kit (Thermo Fisher Scientific, Waltham, MA, USA) on semi-automated Agilent™ Bravo liquid handlers (Agilent Technologies, Santa Clara, CA, USA). Briefly, each well in a 1 mL 96-well plate is prepared with 5 µL proteinase K, 200 µL sample, 275 µL lysis buffer/binding beads, and 5 µL MS2 phage control. It is then shaken for 2 minutes at 1,050 rpm and incubated for 5 minutes at 65°C. Incubation on a magnetic plate at room temperature allows aspiration of waste material and a sample undergoes three cycles of resuspension/aspiration in 165 µL wash buffer, 165 µL 80% ethanol, and 50 µL elution buffer, respectively. After final separation event, 50 µL of purified RNA solution is transferred to a fresh 1 ml 96-well plate.

RT-qPCR was performed according to the U.S. FDA emergency use authorization (EUA) instructions for use (IFU) for TaqPath™ COVID-19 Combo kit (Thermo Fisher Scientific, Waltham, MA, USA), which is a multiplex assay to detect SARS-CoV-2 open reading frame (ORF) 1ab, nucleocapsid (N), and spike (S) genes, in addition to a spiked-in MS2 phage extraction control (reporter dyes FAM, VIC, ABY, and JUN, respectively). The sample volume of 25 µL involves 10 µL of purified RNA and 15 µL of reaction mix that includes TaqPath™ 1-Step Multiplex Master Mix (No ROX™), primers, and probes. Thermal profile parameters involve a 2-minute UNG incubation cycle at 25°C, a 10-minute reverse transcriptase incubation cycle at 53°C, a 2-minute activation cycle at 95°C, followed by 40 cycles of a 3-second denaturation at 95°C and 30-second anneal/extension cycle at 60°C on Applied Biosystems™ 7500 Fast Dx Real-Time PCR Instrument (Thermo Fisher Scientific, Waltham, MA, USA).

Relative genome copy equivalents (GCE) were calculated via averaged standard curves using all (16) 7500 Fast Dx RT-PCR instruments in house, whereby threshold cycle (C_T_) measurements for 5000, 1000, 500, 250, 100, 50, 10, and 5 gce/µL were evaluated for three lots of TaqPath™ COVID-19 Positive Control.

### Matrix Effect Study

The C_T_ values for 6 moderate-to-weakly positive samples [ORF1ab:26.19±4.68; N:27.15±4.35; S:26.61±5.06] were compared in parallel dilution series of 1:2, 1:4, 1:8, 1:16, 1:32, and 1:64 using either VTM (standard) or Negative Sample Pool (matrix), prepared from 16 confirmed negative samples, as diluent. Results were converted to a Matrix Effect factor using the formula: *ME* = −(((*Matrix C*_*T*_/*Standard C*_*T*_) * 100) − 100), whereby a factor of 100 is subtracted from the percent ratio of results to produce a normalized value indicating the magnitude of the suppression or enhancement imparted by the matrix diluent. To correct for the inverse relationship of C_T_ value to concentration, the equation was negated; therefore, a negative ME value indicates suppression, while a positive ME value indicates enhancement.

### Pooled Probit Analysis

Replicates of moderate-to-weak positive sample pools [12 – ORF1ab:27.76±0.82, N:28.85±0.18, S:28.28±0.54 C_T_; 8 – ORF1ab:27.08±0.22, N:28.07±0.25, S:27.41±0.34 C_T_] were produced in two series dilution assays using negative sample pool material as the diluent. The first involved 12 replicates ranging from neat to 1:2, 1:4, 1:8, 1:16, 1:32, 1:64, 1:128, and 1:256 dilutions while the second involved 8 replicates ranging from neat to 1:10, 1:20, 1:40, 1:80, 1:160, 1:320, 1:640, and 1:1280 dilutions; negative sample pools additionally assayed neat separately to rule out presence of interfering substances. Results that met the TaqPath™ method qualitative criteria of <37 C_T_ were counted in the proportion of success rate and the average C_T_ value for each gene of those results were converted to a relative GCE per milliliter. The Probit of the success rate was produced with Excel® formula 5+NORMSINV(P) and was plotted against the log_10_ of the GCE. The 95% confidence interval was calculated from the linear regression analysis to evaluate the limit of detection (LoD).

### 1:5 Dilution Trials

Thirty-four strong-to-weak positive samples [ORF1ab:21.52±5.37; N:22.11±5.36; S:21.77±5.71] were assayed neat and at 1:5 dilution, in singlet, using negative sample material as the diluent to determine a predictable shift in C_T_ value. The neat sample was extracted from 200 µL and the 1:5 dilution was extracted from 40 µL of positive sample diluted in 160 µL of negative sample material.

## Results

### Demographics

Since LSTC establishment in July 2020, the site has performed over 500,000 tests (ca. January 2021) with an overall positivity rate of 0.16%. Cohort descriptions for the positive sample repository used in this study included symptomatic (19.6%; 44% male, 56% female) and asymptomatic (80.4%; 55% male, 45% female) patients within their respective age ranges: 18-24 (63%), 25-31 (17%), 32-49 (13%), and >50 (7%).

### Matrix Effect Study

Of the 77 data-points generated, 31 trials (40.3%) demonstrated signal suppression by the negative sample matrix, whereas 46 trials (59.7%) demonstrated an enhanced signal. Focusing on the range of 2 to 8 pooled samples, a more equally distributed effect is observed with 48% of 46 datapoints showing average suppression of -1.64±1.77, -1.32±1.27, and -4.46±6.19 while 52% showed an average enhancement of 3.67±4.32, 1.53±0.89, and 3.28±2.87 for ORF1ab, N and S gene, respectively (Figure 1). Additionally, 37% of the 2-8 sample pools reported a ME within one unit. Thus, with average C_T_ values of 27.6±0.58, 27.9±0.34, and 29.2±0.44, a matrix effect of 1 would elicit a C_T_ value shift of ±0.28, ±0.28, and ±0.29 for ORF1ab, N, and S genes, respectively. Furthermore, only 17% of 2-8 sample pools reported an ME greater than 5, which translates to a C_T_ value shift of >1 cycle.

**Figure 1:**
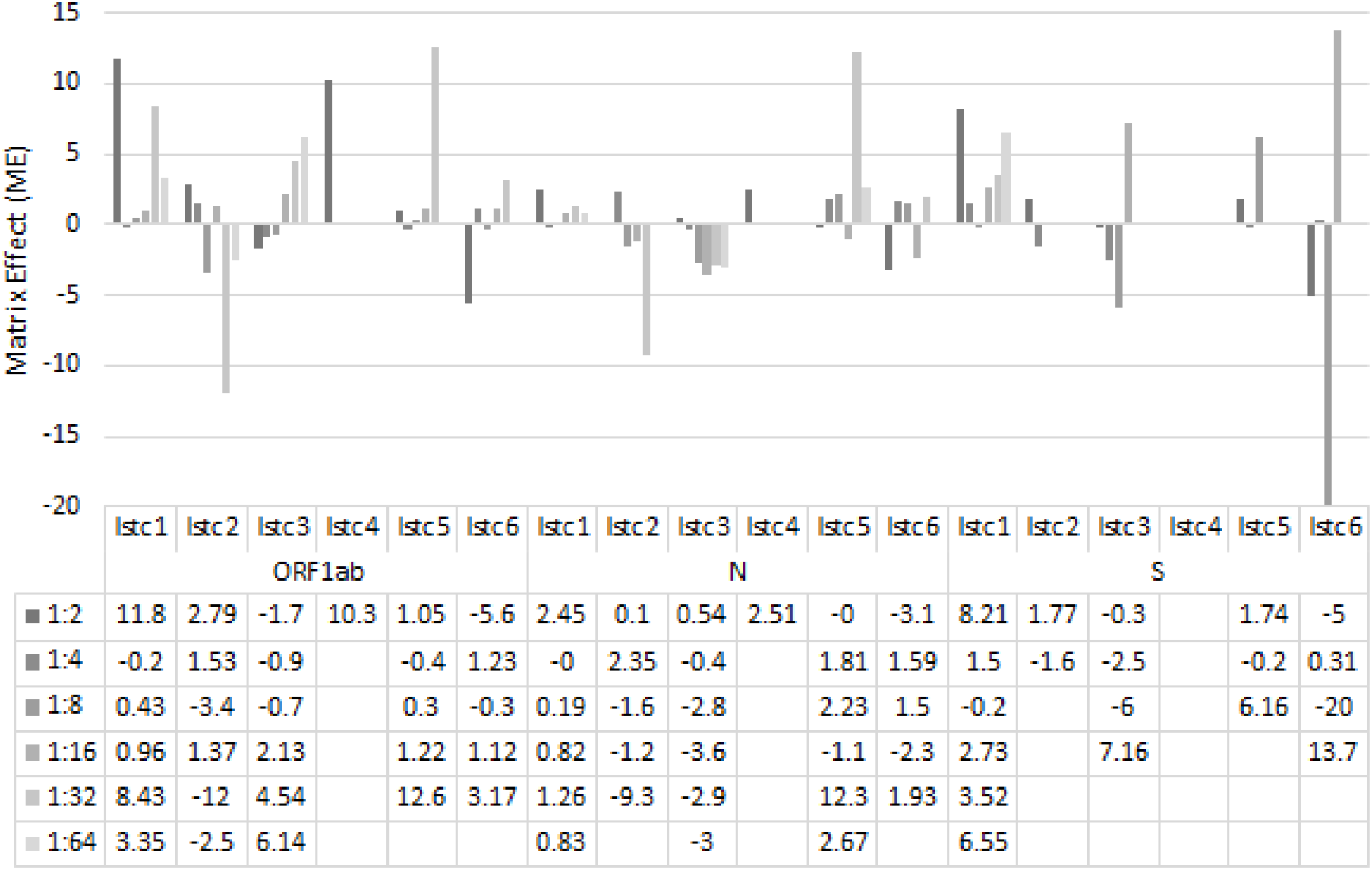
Matrix effect (ME) of diluting positive SARS-CoV-2 sample in negative sample material at 1:2, 1:4, 1:8, 1:16, 1:32, and 1:64 series dilutions. ME factor describes relative enhancing (positive value), or suppressive (negative value) effect of negative sample material on SARS-CoV-2 detection.

### Pooled Probit Analysis

The LOD for each of the SARS-CoV-2 genes using negative sample pool as diluent was 16,075 gce/mL, 1,308 gce/mL, and 1,180,182 gce/mL for the ORF1ab, N, and S genes, respectively. Regressions for the ORF1ab and N gene were more reliable than for the S gene (R^2^ = 0.28), which may be indicative of the reaction mechanism. A qualitative LOD would implement TaqPath™ testing algorithm of 2/3 genes resulted with C_T_ of <37, and LSTC intends to deconvolute a pool with at least 1/3 genes present (Table 1).

**Table 1:** Limit of Detection (LoD) Results of SARS-CoV-2 ORF1ab, N and S gene in negative sample material. Ratio of successes for trials that produced less than 100% hit rate provided with corresponding concentration in genome copy equivalents per milliliter. Slope, y-intercept, and R-squared coefficient of probit analysis used to achieve limit of detection.

| Analyte | Replicates detected at respective GCE/mL |  |  |  | slope | y-int | R<sup>2</sup> | Probit (95% CI) GCE/mL |
| --- | --- | --- | --- | --- | --- | --- | --- | --- |
| ORF1ab | 1495 | 995 | 280 | 163 | 1.24 | 1.43 | 0.91 | 16075 |
|  | [8/12] | [4/8] | [3/8] | [2/12] |  |  |  |  |
| N | 823 | 447 | 298 | 148 | 2.71 | -1.79 | 0.82 | 1308 |
|  | [7/8] | [6/8] | [3/12] | [2/8] |  |  |  |  |
| S | - | 3733 | 2147 | 425 | 0.72 | 2.27 | 0.28 | 1180182 |
|  | - | [5/8] | [2/12] | [2/8] |  |  |  |  |

### 1:5 Dilution Trials

Diluting positive SARS-CoV-2 sample in negative sample material at a 1:5 dilution produced an average shift of the C_T_ value by 2.31±1.16, 2.23±1.12, and 2.79±1.40 cycles for ORF1ab, N, and S gene, respectively (Figure 2). Paired t-test analysis comparing neat and diluted groups for each gene showed no statistically significant difference, with p-values of 0.0878, 0.0988, and 0.0527 for OFR1ab, N, and S gene, respectively. Consistent with the matrix effect study and probit analysis, the S gene exhibited the least sensitivity with 12% of trials losing signal (Figure 2C). Nevertheless, each trial met TaqPath™ testing algorithm for 2/3 genes testing positive to require deconvolution.

**Figure 2:**
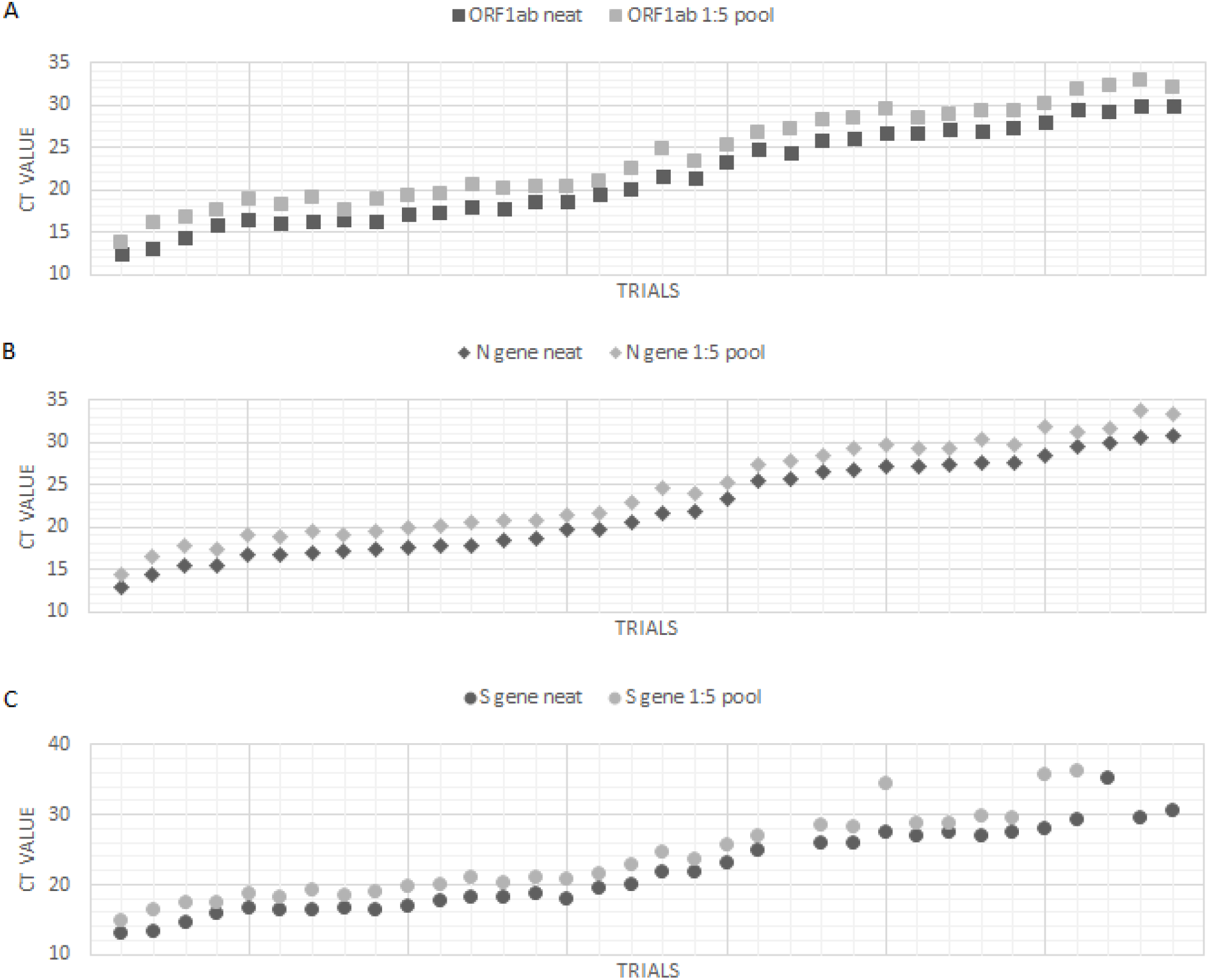
Dilution trials comparing neat to 1:5 dilution of positive SARS-CoV-2 sample in negative patient material. Cycle threshold (C_T_) values for ORF1ab (**A**), N (**B**), and S (**C**) gene observed for each trial (p>0.05).

## Discussion

This study has established an optimal pool size of five specimens to provide a cost-effective bandwidth with respect to prevalence and rate of repeat testing while insignificantly impacting the analytical sensitivity for the Thermo Fisher™ TaqPath™ COVID-19 Combo Kit.

The pandemic has disproportionately affected the mission of IHE with an impact ranging from increased maintenance costs to the academic performance of students. Pre-symptomatic COVID-19 infections are credited with a greater rate of transmission than asymptomatic infections and a correlation with age has been observed with younger populations demonstrating higher rates of asymptomatic cases [16,17]. In the Northeastern University community 80.4% of positive cases from August 2019 to January 2021 were asymptomatic individuals, of which 76% were between the age of 18-27; this data is consistent with results from the University of Georgia [18]. The Northeastern University student community is required to test every three days (+/- 1 day) and employees on site five days a week are expected to test at least twice a week.

Under non-pooling conditions, the sum of assays informed by the MS2 control could be expressed as *E* = *n* + 1, where *E* is the expected number of tests and *n* is the probability of repeat testing due to absence of MS2 signal. Pursuant to LSTC protocol, an MS2 failure is repeated once and further representation for the total assays could be expressed as *E* = (*n* + 1) + (*n*^2). Our modeling indicates the number of repeated testing due to MS2 failure is higher at lower group sizes. It may serve as the deciding factor at a prevalence upwards of 1% since labor costs may outweigh a less than 50% reduction in materials (Figure 3). Furthermore, group testing is an environmentally conservative measure that aligns with ‘green laboratory’ efforts by minimizing the requirements for non-biodegradable materials, toxic chemical waste, and energy consumption [19]. Additional stage hierarchical and array methods may also increase group testing efficiency for diseases of low prevalence [20].

**Figure 3:**
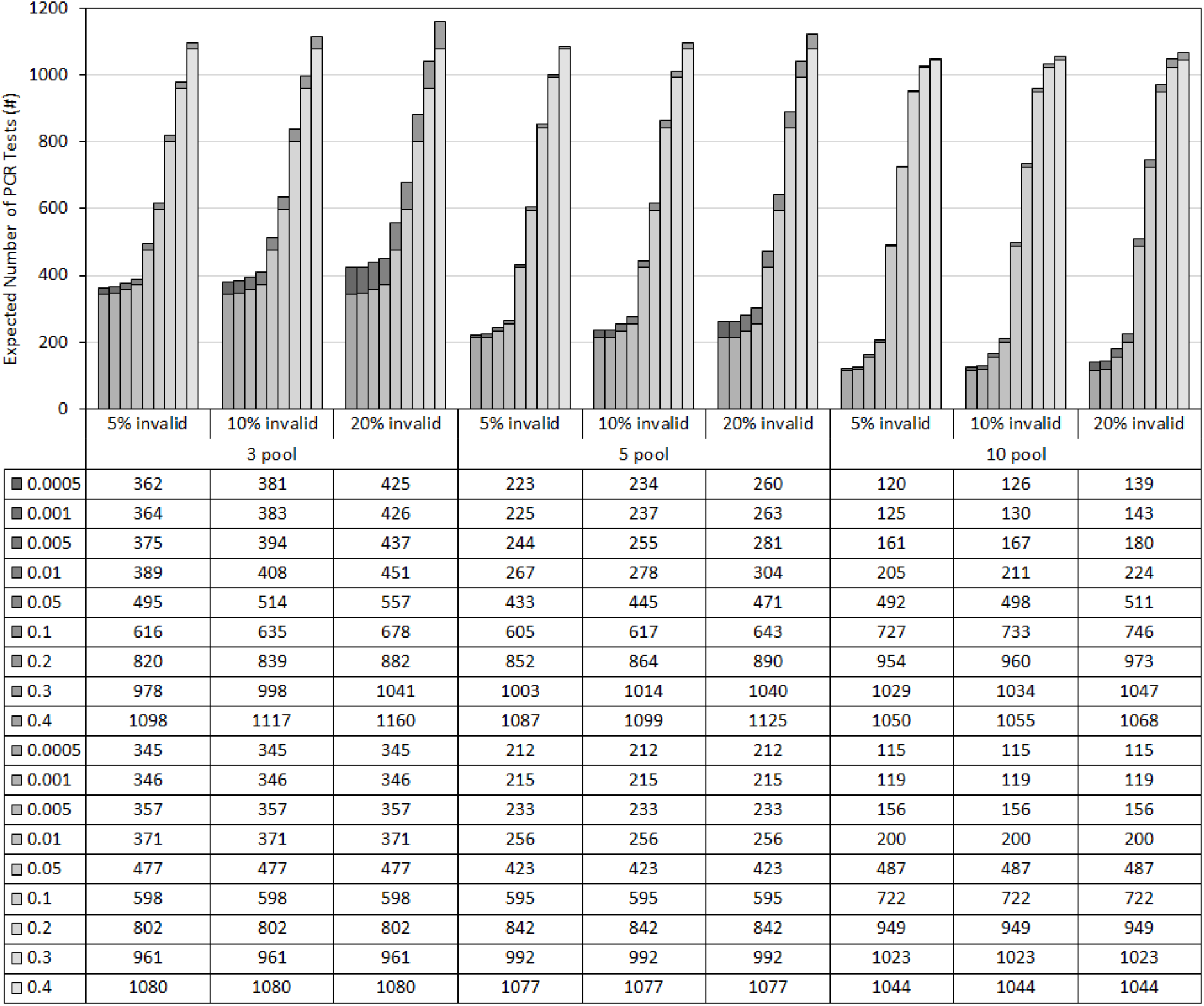
Number of PCR reactions required for testing 1,000 patient samples with respect to pool size, prevalence rate. Dark grey prevalence rates: predicted number of tests including the invalid rate (n). Light grey prevalence rates: predicted number of tests without inclusion of invalid rate.

Multiplex RT-qPCR has been sought to increase assay sensitivity for the detection of SARS-CoV-2, yet inherent limitations arise from increased competition in reaction kinetics [21]. The genomic architecture of SARS-CoV-2 exhibits extensive secondary structure with regions consisting of high base pairing content thought to be mechanistically advantageous for the transcription and thermodynamic stability of sub-genomic RNA [22]. Further consideration that retro-transcription occurs from the 3’ direction suggests cDNA production from the N gene with less deviation than the S and ORF1ab genes. Concurrent with our results, the N gene demonstrated the greatest stability within the negative sample matrix, which was confirmed by a limit of detection 1 and 3 orders of magnitude less than the ORF1ab and S genes, respectively. Recent identification of a SARS-CoV-2 variant of concern (VOC) 202012/01 has been associated with several mutations in the S gene including a deletion at position 69 and 70 to cause S-gene target failure by the TaqPath COVID-19 method [23]. Due to the enhanced binding interactions of VOC 202012/01 spike protein with the target angiotensin converting enzyme 2 (ACE-2), estimates suggest the increased transmissibility will permit strain predominance in the United States by spring 2021 [24]. While no genomic sequence data was available for the samples used in this study, it is important to consider the international nature of the university population as many Northeastern students are from abroad. Accurate assessment of the origins of predominant strains is further challenged by the transience of personal interactions in urban environments. It is also notable that Northeastern campuses in Boston, Massachusetts and Portland, Maine experience significant in-state commuter traffic.

Results from this retrospective study are consistent with recent reports confirming an insignificant effect on the false negative rate for pools of similar size; additionally, our average C_T_ value shifts were precise [14,15]. Probit analysis is applicable to the bimodal output of RT-qPCR for LoD analysis. As all constituent samples of positive pools initially tested positive for all three genes, the data supports a mechanistic basis for the substantial reduction in S-gene sensitivity, which may align with current S-gene target failure hypotheses [23,24]. Our report pioneered a quantification of the matrix effect for RT-qPCR to improve analyte stability. This study was not able to comment on the logistical impacts of deconvolution, which could potentially prolong the <48-hour turn-around-time regularly scheduled testing may require, yet the evidence presented in this study supports pooling as a means of addressing supply chain restrictions. Aliquoting samples to 96-well plates prior to pooling limits operator errors and improved workflow of maintaining operator audit logs and having a well-functioning laboratory information management system (LIMS) program enables easy results review.

## Data Availability

Data can be made available upon request.

## Conflict of Interest

The authors declare no conflict of interest.

## Ethical Statement

Retrospective samples were deidentified for study purposes in accordance with IRB approval by the Northeastern University Office of Human Subject Research Protection (HSRP).

## Acknowledgements

The authors would like to thank Dr. Christopher Bilder for accommodating our inquiries as well as acknowledge the tireless efforts of all at the LSTC.

## Notes

### Competing Interest Statement

The authors have declared no competing interest.

### Funding Statement

No funding statement

### Author Declarations

Northeastern University's IRB provide reviewed and provided and exemption letter for this research

